# ACE-2 Expression in the Small Airway Epithelia of Smokers and COPD Patients: Implications for COVID-19

**DOI:** 10.1101/2020.03.18.20038455

**Authors:** Janice M. Leung, Chen X. Yang, Anthony Tam, Tawimas Shaipanich, Tillie-Louise Hackett, Gurpreet K. Singhera, Delbert R. Dorscheid, Don D. Sin

## Abstract

**Introduction:** Coronavirus disease 2019 (COVID-19) is a respiratory infection caused by the severe acute respiratory syndrome coronavirus-2 (SARS-CoV-2). This virus uses the angiotensin converting enzyme II (ACE-2) as the cellular entry receptor to infect the lower respiratory tract. Because individuals with chronic obstructive pulmonary disease (COPD) are at increased risk of severe COVID-19, we determined whether ACE-2 expression in the lower airways was related to COPD and cigarette smoking.

**Methods:** Using RNA-seq, we determined gene expression levels in bronchial epithelia obtained from cytologic brushings of 6^th^ to 8^th^ generation airways in individuals with and without COPD. We eternally validated these results from two additional independent cohorts, which used microarray technologies to measure gene expression levels from 6^th^ to 12^th^ generation airways.

**Results:** In the discovery cohort (n=42 participants), we found that ACE-2 expression levels were increased by 48% in the airways of COPD compared with non-COPD subjects (COPD=2.52±0.66 log2 counts per million reads (CPM) versus non-COPD= 1.70±0.51 CPM, p=7.62×10^−4^). There was a significant inverse relationship between ACE-2 gene expression and FEV1% of predicted (r=-0.24; p=0.035). Current smoking also significantly increased ACE-2 expression levels compared with never smokers (never current smokers=2.77±0.91 CPM versus smokers=1.78±0.39 CPM, p=0.024). These findings were replicated in the two eternal cohorts.

**Conclusions:** ACE-2 expression in lower airways is increased in patients with COPD and with current smoking. These data suggest that these two subgroups are at increased risk of serious COVID-19 infection and highlight the importance of smoking cessation in reducing the risk.

---

The World Health Organization (WHO) has declared coronavirus disease 2019 (COVID-19) as a pandemic [1]. COVID-19 is caused by severe acute respiratory syndrome coronavirus-2 (SARS-CoV-2). COVID-19 displays symptoms ranging from mild to severe (pneumonia) that can lead to death in some individuals [2-4]. As of March 13, 2020, there have been 144,064 laboratory-confirmed cases of COVID-19 worldwide with 5,397 deaths worldwide[5]. SARS-CoV-2 uses the angiotensin converting enzyme II (ACE-2) as the cellular entry receptor[6]. While the virus can infect individuals of any age, to date, most of the severe cases have been described in those over the age of 55 years and with significant co-morbidities such as chronic obstructive pulmonary disease (COPD) [7]. Here, we determined whether patients with COPD have increased expression of ACE-2 in bronchial epithelial cells in lower respiratory tract.

Patients undergoing bronchoscopy at St. Paul’s Hospital (SPH), Vancouver, Canada for clinical purposes were enrolled (Table 1). The protocol was approved by the University of British Columbia/Providence Health Care Ethics Board (UBC/PHC REB H15-02166). All patients were required to be 19 years of age or older, who underwent spirometry according to international guidelines[8]. Patients with COPD were defined as those having a clinical diagnosis of COPD made by a board-certified respiratory physician and either a forced expiratory volume in 1 second (FEV1)/forced vital capacity (FVC) <70% or a clear evidence of emphysema on computed tomographic (CT) imaging on visual inspection. Cytologic brushings were obtained in subsegmental airways (6^th^-8^th^ generation) of the lung that were unaffected by the patient’s underlying clinical indication for bronchoscopy.

**Table 1.**
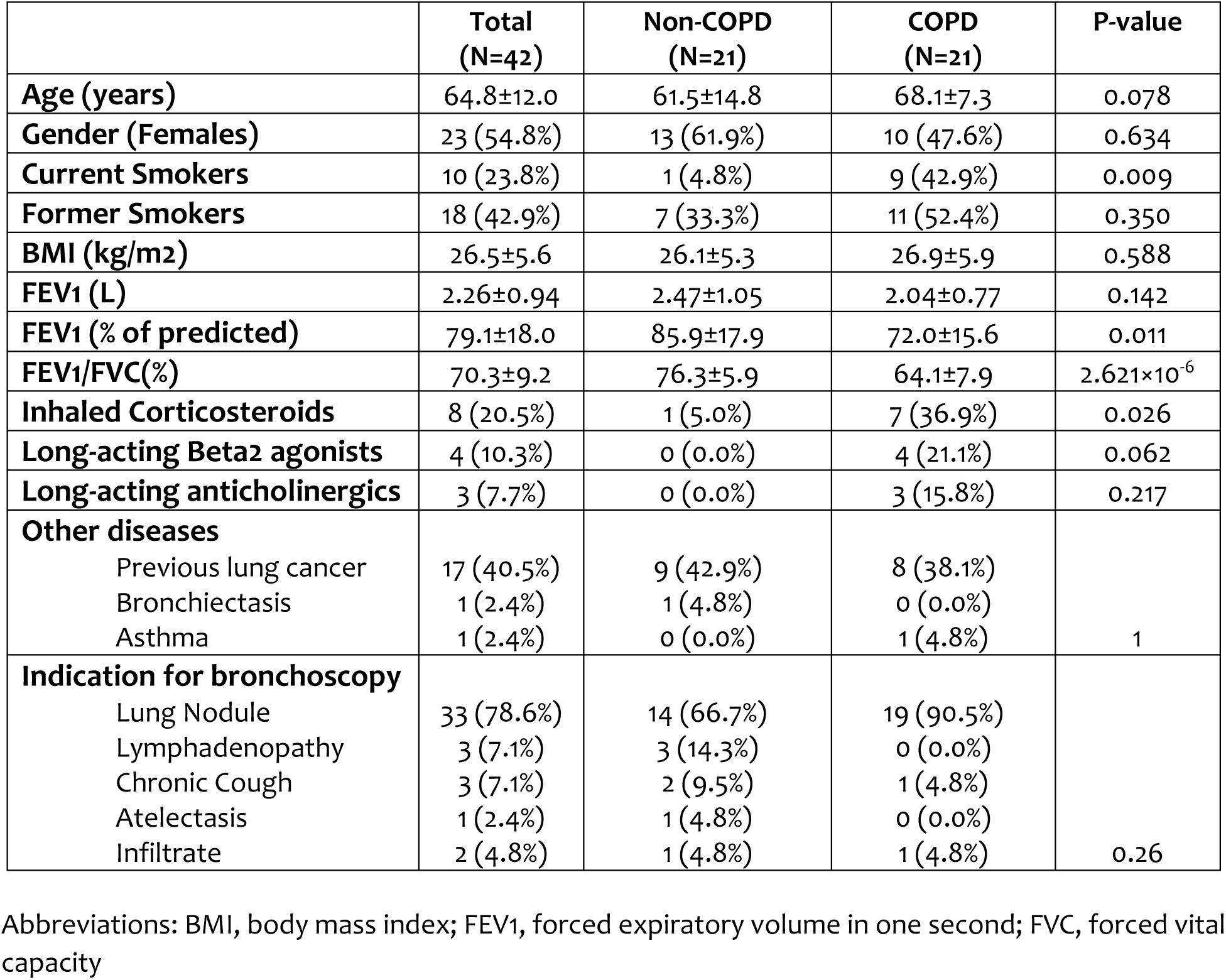
Demographics and Clinical Features of St. Paul’s Hospital (SPH) Cohort. Continuous data are expressed as mean±SD; and dichotomous data as number (% of column totals) P-values were obtained using Student’s t-test or Fisher’s exact test where appropriate.

Total RNA was extracted from cytologic brushings using the RNeasy Mini Kit (Qiagen, Hilden, Germany). Transcriptome sequencing was performed on the NovaSeq 6000 (Illumina, San Diego, CA) at a sequencing depth of 55 million reads. Raw sequencing reads were quality controlled with FastQC[9] and aligned to the GENCODE (version 31) GRCh37 genome reference using STAR (Spliced Transcripts Alignment to a Reference) [10]. After alignment, the data were quantified using RSEM (RNA-Seq by Expectation Maximization) to obtain the read counts. Limma voom[11] was applied to normalize the counts to log2 counts per million reads (CPM), which was used in the downstream analysis.

Two cohorts were used for validation; the details of which are provided in a previous publication [12]. First, we used 16 datasets obtained from bronchial brushings of 10^th^-12^th^ generation bronchi collected at a single center using the U133 Plus 2.0 microarray (denoted as the Cornell Dataset)[13]. Second, we used dataset GSE37147 consisting of bronchial brushings from the 6^th^-8^th^ generation airways with gene expression profiles generated from the GeneChip Human Gene 1.0 ST microarray[14]. This dataset was denoted as British Columbia Cancer Agency (BCCA) cohort.

We also determined protein expression of ACE-2 in resected lung tissue specimens. These samples were obtained from 10 current smokers with COPD (mean±SD, FEV1/FVC 61±7%), 9 non-smoker controls (FEV1/FVC 85±2%), and 8 healthy current smokers (FEV1/FVC 78±6%). Human lung tissue samples were obtained with informed consent from patients undergoing thoracic surgery as part of the James Hogg Lung Registry (UBC/PHC REB Protocol H00-50110). Formalin-fixed paraffin-embedded human lung tissues were stained with antibody against ACE-2 (Ab15348; Abcam) using the Bond Polymer Refine Red Detection kit on the Leica Bond Autostainer as previously described[15]. Airway epithelial-specific ACE-2 protein intensity was quantified using the Aperio imaging system with normalization to the length of the basement membrane (Leica Biosystem; Concord, Ontario).

For the primary study population, log2 CPM of ACE-2 was the principal outcome of interest. Robust linear models were used to determine whether 1) ACE-2 was differentially expressed in patients with COPD and in smokers after adjustment for age and sex and 2) ACE-2 expression was significantly correlated with lung function. All analyses were performed in R (version 3.5.0). In the immunohistochemistry dataset, Kruskal-Wallis test with Dunn’s Multiple Comparisons test was used.

Table 1 displays the demographic and clinical characteristics of the SPH cohort. ACE-2 expression in the epithelial cells was significantly increased in COPD versus non-COPD subjects (Mean±SD of non-COPD=1.70±0.51 versus COPD=2.52±0.66, p=7.62×10^−4^; **Figure 1A**). There was a significant inverse relationship between ACE-2 gene expression and FEV1% of predicted (r=-0.24; p=0.035; **Figure 1B)**. Interestingly, smoking status was also significantly related to ACE-2 gene expression levels in airways of these participants with current smokers having a significantly higher gene expression than never smokers (never smokers=1.78±0.39 versus current smokers=2.77±0.91, p=0.024). Former smokers had gene expression levels in-between that of never and current smokers (former smokers=2.00±1.23; **Figure 1C**). Conditional on the smoking status, the association between ACE-2 expression and COPD was still significant (Adjusted Mean±SE of non-COPD: 0.90±0.65 versus COPD: 1.75±0.82, p=0.016).

**Figure 1A:**
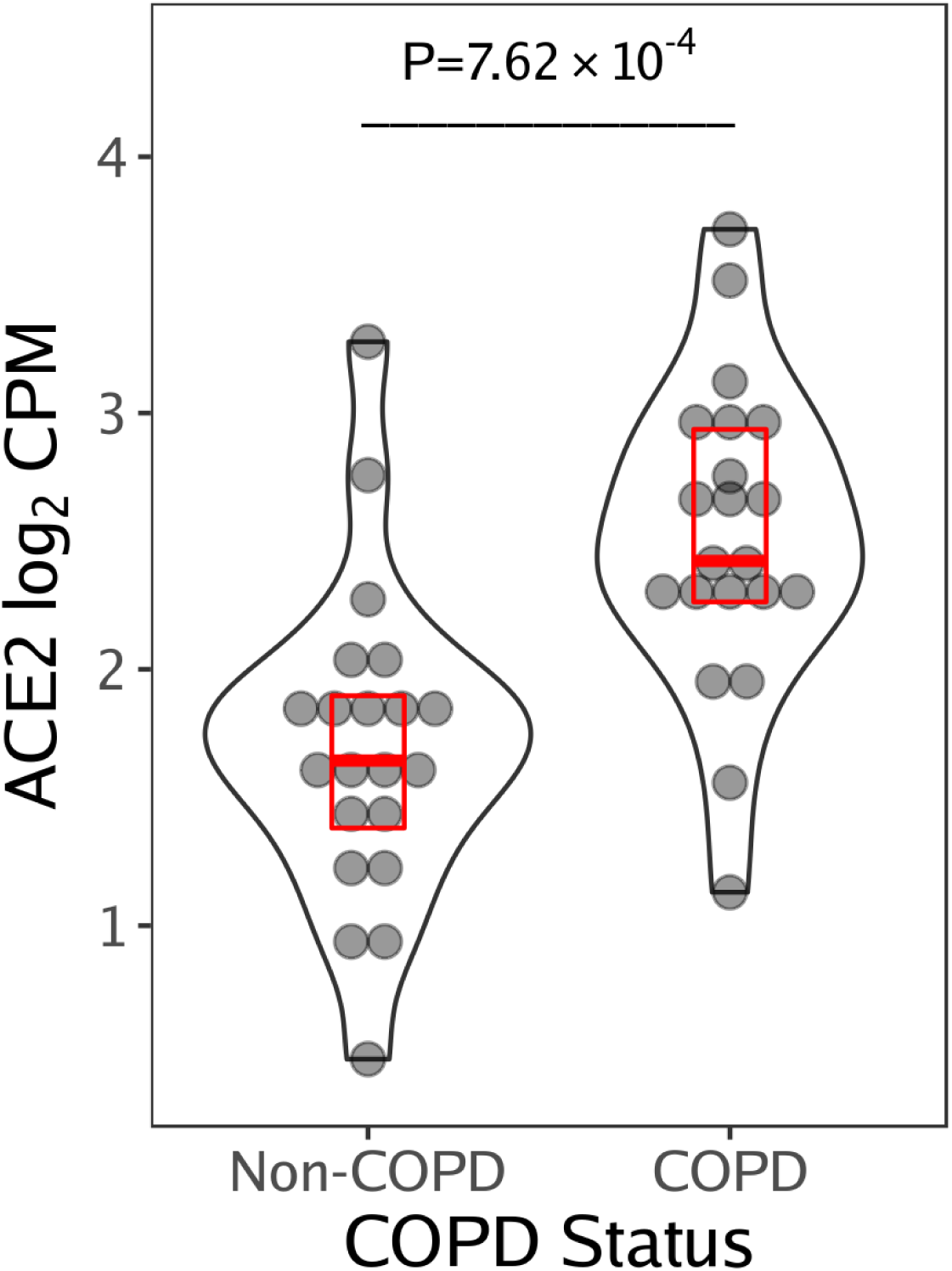
A violin plot of ACE-2 expression in small airways of COPD and non-COPD subjects in the St. Paul’s Hospital Cohort. The red box indicates the median and the interquartile range. The P-value was obtained from the robust linear model Abbreviations: ACE-2, angiotensin converting enzyme II; COPD, chronic obstructive pulmonary disease; CPM, counts per million reads

**Figure 1B:**
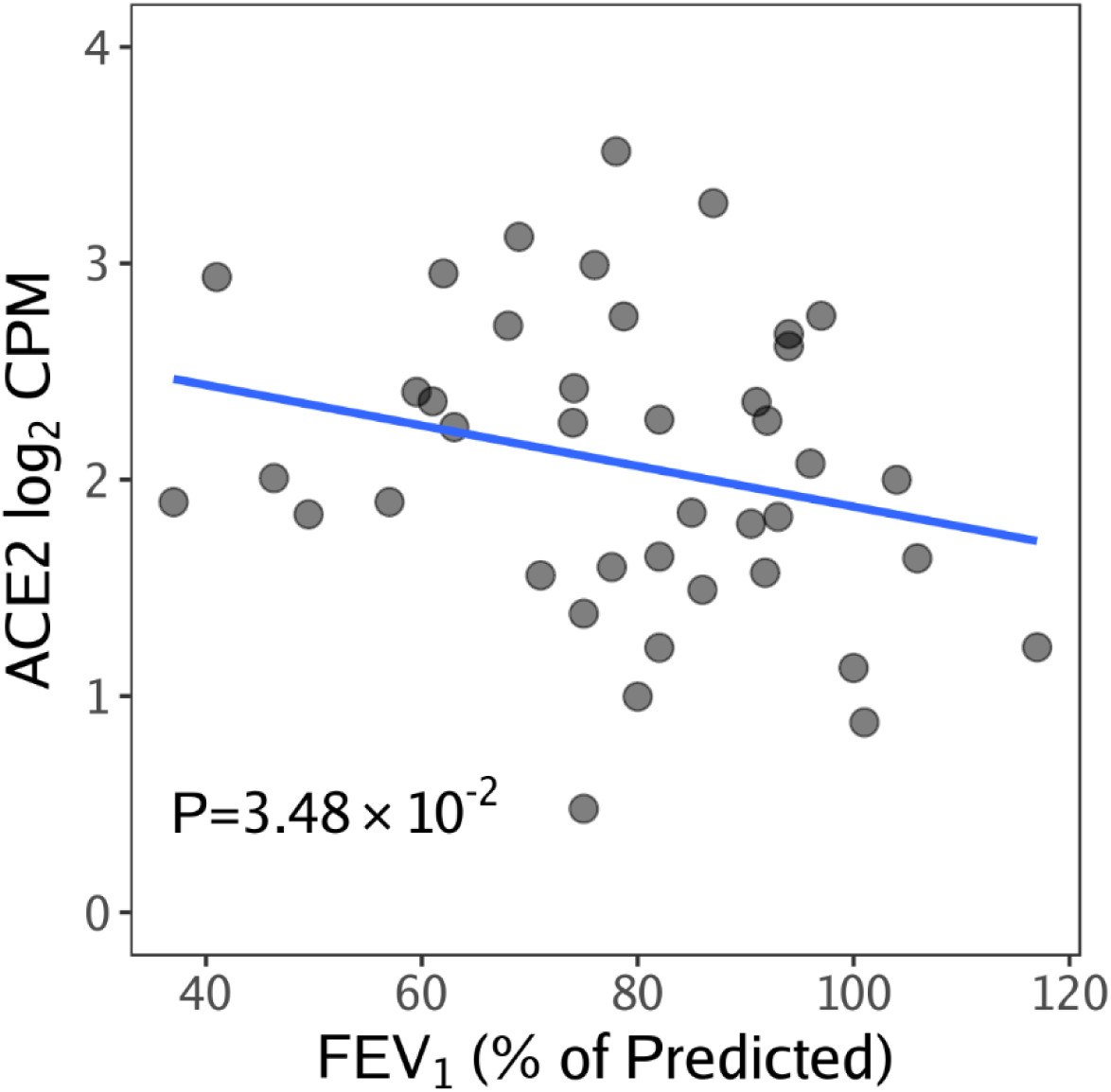
A scatter plot of ACE-2 expression in small airways according to FEV1 % Predicted in the St. Paul’s Hospital Cohort. Abbreviations: ACE-2, angiotensin converting enzyme II; COPD, chronic obstructive pulmonary disease; CPM, counts per million reads ACE-2 gene expression in airway epithelia is inversely related to FEV1% predicted (p=0.0348)

**Figure 1C.**
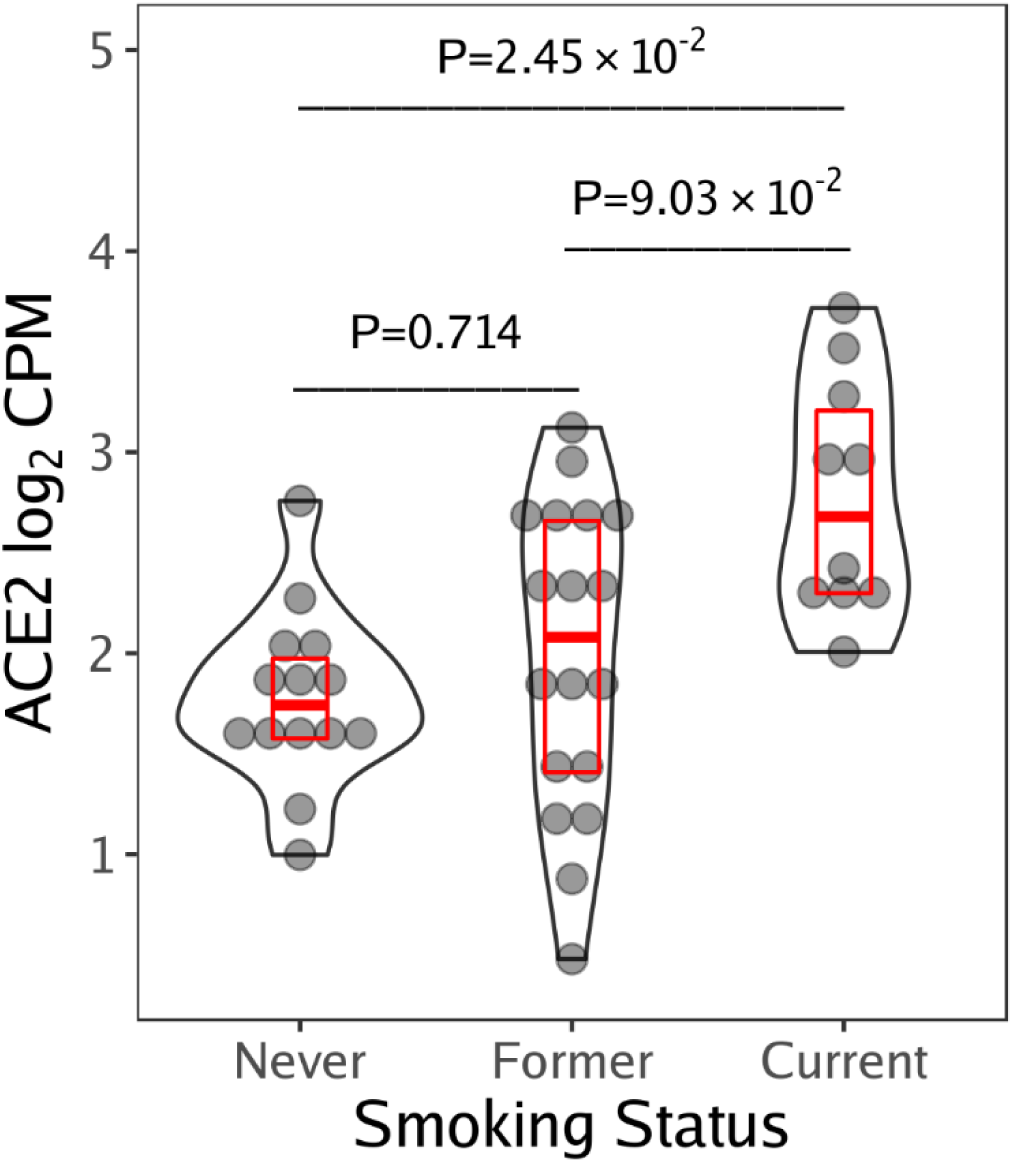
A violin plot of ACE2 expression in small airways of never, former and current smokers in the St. Paul’s Hospital Cohort. The red box indicates the median and the interquartile range. The P-value was obtained from the robust linear model. Abbreviations: ACE-2, angiotensin converting enzyme II; CPM, counts per million reads

Next, we validated the above findings in: 1) the Cornell Cohort (N= 211) and 2) British Columbia Cancer Research Agency (BCCA) cohort (N=238). The average age of the Cornell Cohort was 43.6 (SD=10.5) years with 33.2% of the cohort being females. There were 32.2% who were never smokers and 67.8% who were current smokers at the time of the bronchoscopy. The average age of the BCCA cohort was 64.5 (SD=5.9) years with 43.3% of the cohort being females. All were heavy smokers with at least 30 pack-years of smoking. Of these, 41.6% were current smokers at the time of the bronchoscopy and the remaining were former smokers.

In both the Cornell and BCCA cohorts, current smokers had increased ACE-2 gene expression levels in the airways compared with never smokers (in the Cornell cohort; Mean±SD of never smokers=4.15±0.36 versus current smokers=4.34±0.45, p=1.92×10^−3^) and with former smokers (in the BCCA cohort; Mean±SD of former smokers=5.57±0.37 versus current smokers=6.05±0.53, p<2×10^−16^). In the BCCA cohort, pre-bronchodilator FEV1 was measured and it was significantly related to ACE-2 gene expression level (r=-0.10;p=0.037).

Representative images of epithelial-specific ACE2 protein expression in non-smokers, healthy smokers and smokers with COPD are shown in Figure 1D. ACE-2 expression in the human small airway epithelium was significantly increased in COPD compared to non-smokers but not in healthy smokers (Figure 1D).

**Figure 1D.**
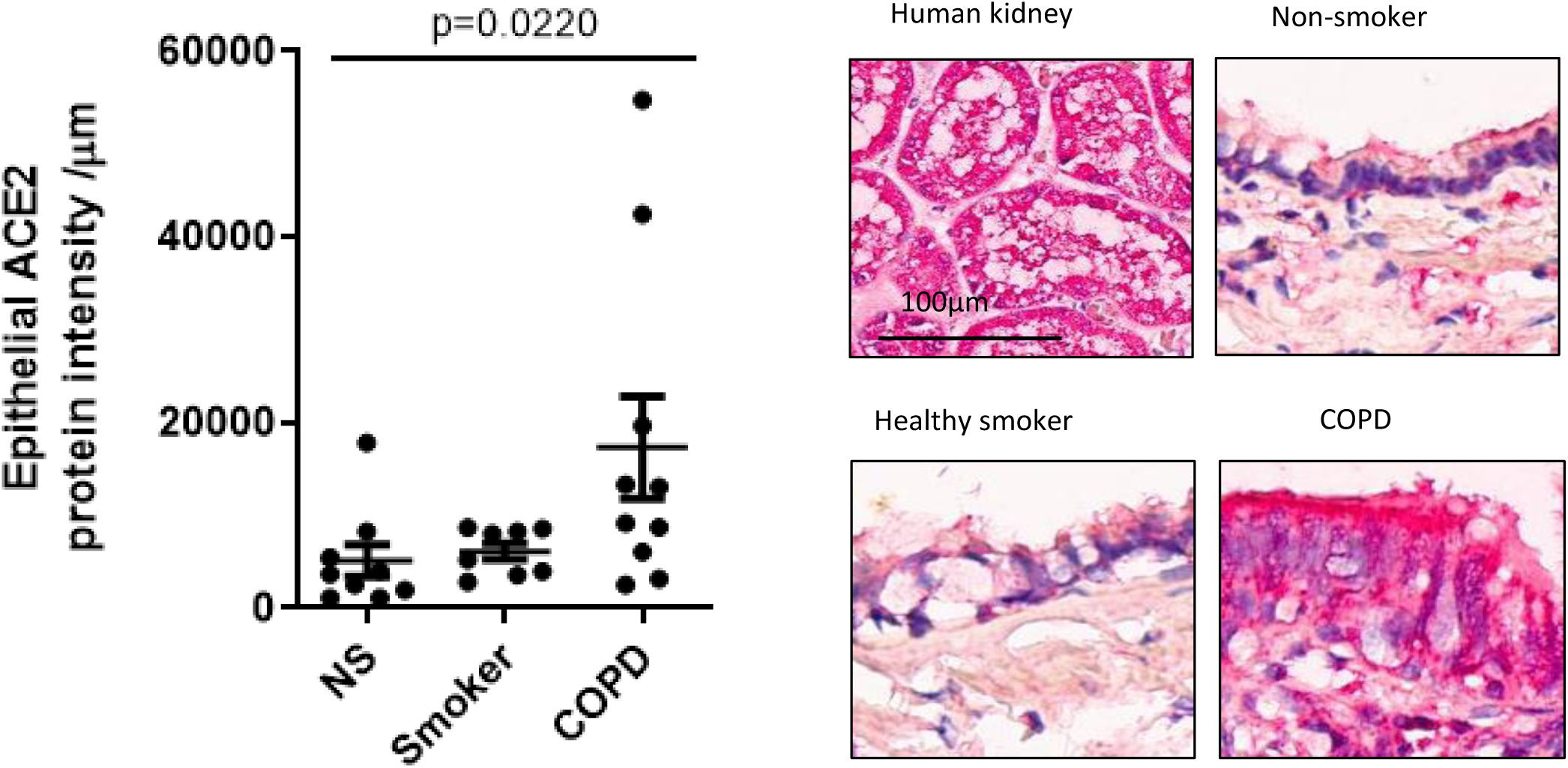
Protein staining of ACE2 airways of Individuals with and without COPD. Human kidney slide was the positive control for ACE-2. In airways, most of the protein expression was noted in the epithelial layer, most pronounced in those with COPD. Abbreviations: ACE-2, angiotensin converting enzyme II; NS, non-smoker

There is a worldwide outbreak of COVID-19 coronavirus. Although most patients infected and diagnosed with CVOID-19 disease have mild symptoms, approximately 20% of individuals have demonstrated severe or critically severe disease including symptoms and signs of pneumonia, respiratory failure, septic shock and multi-organ failure. The estimated case-fatality rate is 1-2% [2, 3]. Importantly, nearly all deaths have occurred in those with significant underlying chronic diseases including COPD, and cardiovascular diseases [4]. The reason for this observation is largely unknown.

One possibility is differential expression of the ACE-2, which is the main receptor used by SARS-CoV-2 to gain entry into the host mucosa and cause active infection. Here, we investigated gene expression levels of ACE-2 in the airways of individuals with and without COPD and found that COPD and current smokers had significantly increased expression of ACE-2. Importantly, gene expression levels of ACE-2 were inversely related to individual’s FEV1, suggesting a dose-dependent response. These findings were observed in 3 different cohorts, indicating their generalizability and robustness.

ACE-2 is a type I transmembrane metallocarboxypeptidase with homology to angiotensin converting enzyme (ACE). In contrast to ACE, which converts angiotensin I to the active vasoconstrictor, angiotensin II, ACE-2 breaks down angiotensin II to its metabolites including angiotensin-(1–9) and angiotensin-(1–7), which are potent vasodilators, and thus may be a negative regulator of the renin-angiotensin system[16]. ACE-2 is expressed in a variety of different tissues including both the upper and lower respiratory tract, myocardium and the gastrointestinal mucosa [17]. Although its role in human health and disease has not been fully elucidated, it appears to have an important regulatory role in blood pressure and cardiac function. The physiologic role of ACE-2 in the airways is largely unknown. However, in mice, ACE-2 has been shown to protect animals from severe lung injury related to aspiration and sepsis [18].

To our knowledge, our study is the first to demonstrate increased ACE-2 expression in airways of current (but not former) smokers and those with COPD. These results are also consistent with previous observations in small animals wherein smoke exposure has been shown to upregulate both the expression and activity of ACE-2 in the airways [19, 20]. While the up-regulation of ACE-2 may be useful in protecting the host against acute lung injury, chronically, this may predispose individuals to increased risk of coronavirus infections, which uses this receptor to gain entrance into epithelial cells. This may in part explain the increased risk of viral respiratory tract infection in active smokers and virus-related exacerbations in those with COPD.

There were limitations to the study. First, the study was cross-sectional and as such, we could not determine whether interventions such as inhaled corticosteroids or bronchodilators (for those with COPD) could modulate ACE-2 gene expression in the airways. Second, as receptor expression is one of many host factors that govern infection risk among individuals, the precise attributable risk (for coronavirus infections) imposed by cigarette smoking and COPD is uncertain. Third, although the airway epithelia is the major source of entry for COVID-19, the virus can gain host entry through other ports including gastrointestinal mucosa, which was not evaluated in this study. Fourth, we did not have access to upper airway tissues, which may also become infected with SARS-CoV-2.

In summary, active cigarette smoking and COPD up-regulate ACE-2 expression in lower airways, which in part may explain the increased risk of severe COVID-19 in these sub-populations. These findings highlight the importance of smoking cessation for these individuals and increased surveillance of these risk subgroups for prevention and rapid diagnosis of this potentially deadly disease.

## Data Availability

Data are available on GEO or upon request from senior author (DDS)

## References

1. World Health Organization. Coronavirus disease (COVID-19) outbreak https://www.who.int/emergencies/diseases/novel-coronavirus-2019/events-as-they-happen.

2. Guan W-j, Ni Z-y, Hu Y, Liang W-h, Ou C-q, He J-x, Liu L, Shan H, Lei C-l, Hui DSC, D. B, Li L-j, Zeng G, Yuen K-Y, Chen R-c, Tang C-l, Wang T, Chen P-y, Xiang J, Li S-y, Wang J-l, Liang Z-j, Peng Y-x, Wei L, Liu Y, Hu Y-h, Peng P, Wang J-m, Liu J-y, Chen Z, Li G, Zheng Z-j, Qiu S-q, Luo J, Ye C-j, Zhu S-y, Zhong N-s. Clinical Characteristics of Coronavirus Disease 2019 in China. New England Journal of Medicine 2020.

3. Li Q, Guan X, Wu P, Wang X, Zhou L, Tong Y, Ren R, Leung KSM, Lau EHY, Wong JY, Xing X, Xiang N, Wu Y, Li C, Chen Q, Li D, Liu T, Zhao J, Liu M, Tu W, Chen C, Jin L, Yang R, Wang Q, Zhou S, Wang R, Liu H, Luo Y, Liu Y, Shao G, Li H, Tao Z, Yang Y, Deng Z, Liu B, Ma Z, Zhang Y, Shi G, Lam TTY, Wu JT, Gao GF, Cowling BJ, Yang B, Leung GM, Feng Z. Early Transmission Dynamics in Wuhan, China, of Novel Coronavirus–Infected Pneumonia. New England Journal of Medicine 2020.

4. Wu Z, McGoogan JM. Characteristics of and Important Lessons From the Coronavirus Disease 2019 (COVID-19) Outbreak in China: Summary of a Report of 72?314 Cases From the Chinese Center for Disease Control and Prevention. Jama 2020.

5. Coronavirus Worldometer. available at https://www.worldometers.info/coronavirus/.

6. Zhou P, Yang XL, Wang XG, Hu B, Zhang L, Zhang W, Si HR, Zhu Y, Li B, Huang CL, Chen HD, Chen J, Luo Y, Guo H, Jiang RD, Liu MQ, Chen Y, Shen XR, Wang X, Zheng XS, Zhao K, Chen QJ, Deng F, Liu LL, Yan B, Zhan FX, Wang YY, Xiao GF, Shi ZL. A pneumonia outbreak associated with a new coronavirus of probable bat origin. Nature 2020.

7. Wang D, Hu B, Hu C, Zhu F, Liu X, Zhang J, Wang B, Xiang H, Cheng Z, Xiong Y, Zhao Y, Li Y, Wang X, Peng Z. Clinical Characteristics of 138 Hospitalized Patients With 2019 Novel Coronavirus-Infected Pneumonia in Wuhan, China. Jama 2020.

8. Graham BL, Steenbruggen I, Miller MR, Barjaktarevic IZ, Cooper BG, Hall GL, Hallstrand TS, Kaminsky DA, McCarthy K, McCormack MC, Oropez CE, Rosenfeld M, Stanojevic S, Swanney MP, Thompson BR. Standardization of Spirometry 2019 Update. An Official American Thoracic Society and European Respiratory Society Technical Statement. American journal of respiratory and critical care medicine 2019: 200(8): e70–e88.

9. Andrews S. (2010). FastQC: a quality control tool for high throughput sequence data. available online at: http://www.bioinformatics.babraham.ac.uk/projects/fastqc.

10. Dobin A, Davis CA, Schlesinger F, Drenkow J, Zaleski C, Jha S, Batut P, Chaisson M, Gingeras TR. STAR: ultrafast universal RNA-seq aligner. Bioinformatics 2013: 29(1): 15–21.

11. Law CW, Chen Y, Shi W, Smyth GK. voom: precision weights unlock linear model analysis tools for RNA-seq read counts. Genome Biology 2014: 15(2): R29.

12. Yang CX, Shi H, Ding I, Milne S, Hernandez Cordero AI, Yang CWT, Kim EK, Hackett TL, Leung J, Sin DD, Obeidat M. Widespread Sexual Dimorphism in the Transcriptome of Human Airway Epithelium in Response to Smoking. Sci Rep 2019: 9(1): 17600.

13. Strulovici-Barel Y, Omberg L, O’Mahony M, Gordon C, Hollmann C, Tilley AE, Salit J, Mezey J, Harvey BG, Crystal RG. Threshold of biologic responses of the small airway epithelium to low levels of tobacco smoke. American journal of respiratory and critical care medicine 2010: 182(12): 1524–1532.

14. Steiling K, van den Berge M, Hijazi K, Florido R, Campbell J, Liu G, Xiao J, Zhang X, Duclos G, Drizik E, Si H, Perdomo C, Dumont C, Coxson HO, Alekseyev YO, Sin D, Pare P, Hogg JC, McWilliams A, Hiemstra PS, Sterk PJ, Timens W, Chang JT, Sebastiani P, O’Connor GT, Bild AH, Postma DS, Lam S, Spira A, Lenburg ME. A dynamic bronchial airway gene expression signature of chronic obstructive pulmonary disease and lung function impairment. American journal of respiratory and critical care medicine 2013: 187(9): 933–942.

15. Tam A, Hughes M, McNagny KM, Obeidat M, Hackett TL, Leung JM, Shaipanich T, Dorscheid DR, Singhera GK, Yang CWT, Pare PD, Hogg JC, Nickle D, Sin DD. Hedgehog signaling in the airway epithelium of patients with chronic obstructive pulmonary disease. Sci Rep 2019: 9(1): 3353.

16. Crackower MA, Sarao R, Oudit GY, Yagil C, Kozieradzki I, Scanga SE, Oliveira-dos-Santos AJ, da Costa J, Zhang L, Pei Y, Scholey J, Ferrario CM, Manoukian AS, Chappell MC, Backx PH, Yagil Y, Penninger JM. Angiotensin-converting enzyme 2 is an essential regulator of heart function. Nature 2002: 417(6891): 822–828.

17. Harmer D, Gilbert M, Borman R, Clark KL. Quantitative mRNA expression profiling of ACE 2, a novel homologue of angiotensin converting enzyme. FEBS Lett 2002: 532(1-2): 107–110.

18. Imai Y, Kuba K, Rao S, Huan Y, Guo F, Guan B, Yang P, Sarao R, Wada T, Leong-Poi H, Crackower MA, Fukamizu A, Hui CC, Hein L, Uhlig S, Slutsky AS, Jiang C, Penninger JM. Angiotensin-converting enzyme 2 protects from severe acute lung failure. Nature 2005: 436(7047): 112–116.

19. Hung YH, Hsieh WY, Hsieh JS, Liu FC, Tsai CH, Lu LC, Huang CY, Wu CL, Lin CS. Alternative Roles of STAT3 and MAPK Signaling Pathways in the MMPs Activation and Progression of Lung Injury Induced by Cigarette Smoke Exposure in ACE2 Knockout Mice. Int J Biol Sci 2016: 12(4): 454–465.

20. Yilin Z, Yandong N, Faguang J. Role of angiotensin-converting enzyme (ACE) and ACE2 in a rat model of smoke inhalation induced acute respiratory distress syndrome. Burns 2015: 41(7): 1468–1477.

